# Safety of hydroxychloroquine, alone and in combination with azithromycin, in light of rapid wide-spread use for COVID-19: a multinational, network cohort and self-controlled case series study

**DOI:** 10.1101/2020.04.08.20054551

**Authors:** Jennifer C.E. Lane, James Weaver, Kristin Kostka, Talita Duarte-Salles, Maria Tereza F. Abrahao, Heba Alghoul, Osaid Alser, Thamir M Alshammari, Patricia Biedermann, Edward Burn, Paula Casajust, Mitch Conover, Aedin C. Culhane, Alexander Davydov, Scott L. DuVall, Dmitry Dymshyts, Sergio Fernandez-Bertolin, Kristina Fišter, Jill Hardin, Laura Hester, George Hripcsak, Seamus Kent, Sajan Khosla, Spyros Kolovos, Christophe G. Lambert, Johan van der Lei, Kristine E. Lynch, Rupa Makadia, Andrea V. Margulis, Michael E. Matheny, Paras Mehta, Daniel R. Morales, Henry Morgan-Stewart, Mees Mosseveld, Danielle Newby, Fredrik Nyberg, Anna Ostropolets, Rae Woong Park, Albert Prats-Uribe, Gowtham A. Rao, Christian Reich, Jenna Reps, Peter Rijnbeek, Selva Muthu Kumaran Sathappan, Martijn Schuemie, Sarah Seager, Anthony Sena, Azza Shoaibi, Matthew Spotnitz, Marc A. Suchard, Joel Swerdel, Carmen O. Torre, David Vizcaya, Haini Wen, Marcel de Wilde, Seng Chan You, Lin Zhang, Oleg Zhuk, Patrick Ryan, Daniel Prieto-Alhambra, on behalf of OHDSI-COVID-19 consortium

**Affiliations:** Centre for Statistics in Medicine, NDORMS, University of Oxford; Janssen Research and Development, Titusville, NJ, USA; Real World Solution, IQVIA, Cambridge, MA, USA; Fundació Institut Universitari per a la recerca a l’Atenció Primària de Salut Jordi Gol i Gurina (IDIAPJGol); Faculty of Medicine, University of Sao Paulo, Brazil; Faculty of Medicine, Islamic University of Gaza; Massachusetts General Hospital, Harvard Medical School, Boston, USA; King Saud University, Riyadh, Saudi Arabia; Actelion Pharmaceuticals Ltd, Allschwil, Switzerland; Real-World Evidence, Trial Form Support, Barcelona, Spain; Department of Data Sciences, Dana-Farber Cancer Institute, Department of Biostatistics, Harvard TH Chan School of Public Health, Boston, MA, USA; Medical Ontology solutions, Odysseus Data Services Inc, Cambridge MA; Department of Veterans Affairs, USA; University of Utah School of Medicine, USA; University of Zagreb, School of Medicine, Andrija Štampar School of Public Health; Department of Biomedical Informatics, Columbia University Irving Medical Center, New York, NY, USA; NewYork-Presbyterian Hospital, New York, NY, USA; National Institute for Health and Care Excellence, UK; AstraZeneca, Real World Science & Digital, Cambridge UK; Department of Internal Medicine, University of New Mexico Health Sciences Center, Albuquerque, NM, USA; Erasmus MC, Rotterdam, Netherlands; RTI Health Solutions, Barcelona, Spain; Vanderbilt University, USA; College of Medicine, University of Arizona, USA; Division of Population Health and Genomics, University of Dundee, Scotland, UK.; University of Oxford, Department of Psychiatry, Warneford Hospital, Oxford UK; Institute of Medicine, Sahlgrenska Academy, University of Gothenburg, Gothenburg, Sweden; Department of Biomedical Informatics, Ajou University, Suwon, South Korea; Saw Swee Hock School of Public Health, National University of Singapore, Singapore; Department of Biostatistics, University of California, Los Angeles; Bayer pharmaceuticals, Barcelona, Spain; Shuguang Hospital affiliated to Shanghai University of Traditional Chinese Medicine, Shanghai, China; School of Public Health, Peking Union Medical College, Chinese Academy of Medical Sciences & Melbourne School of Population and Global Health, University of Melbourne

**Keywords:** hydroxychloroquine, chloroquine, covid-19, coronavirus, SARS-CoV-2, safety, epidemiology, international, serious adverse event, rheumatoid arthritis, azithromycin

## Abstract

**Background:** Hydroxychloroquine has recently received Emergency Use Authorization by the FDA and is currently prescribed in combination with azithromycin for COVID-19 pneumonia. We studied the safety of hydroxychloroquine, alone and in combination with azithromycin.

**Methods:** New user cohort studies were conducted including 16 severe adverse events (SAEs). Rheumatoid arthritis patients aged 18+ and initiating hydroxychloroquine were compared to those initiating sulfasalazine and followed up over 30 days. Self-controlled case series (SCCS) were conducted to further establish safety in wider populations. Separately, SAEs associated with hydroxychloroquine-azithromycin (compared to hydroxychloroquine-amoxicillin) were studied. Data comprised 14 sources of claims data or electronic medical records from Germany, Japan, Netherlands, Spain, UK, and USA. Propensity score stratification and calibration using negative control outcomes were used to address confounding. Cox models were fitted to estimate calibrated hazard ratios (CalHRs) according to drug use. Estimates were pooled where I2<40%.

**Results:** Overall, 956,374 and 310,350 users of hydroxychloroquine and sulfasalazine, and 323,122 and 351,956 users of hydroxychloroquine-azithromycin and hydroxychloroquine-amoxicillin were included. No excess risk of SAEs was identified when 30-day hydroxychloroquine and sulfasalazine use were compared. SCCS confirmed these findings. However, when azithromycin was added to hydroxychloroquine, we observed an increased risk of 30-day cardiovascular mortality (CalHR2.19 [1.22-3.94]), chest pain/angina (CalHR 1.15 [95% CI 1.05-1.26]), and heart failure (CalHR 1.22 [95% CI 1.02-1.45])

**Conclusions:** Short-term hydroxychloroquine treatment is safe, but addition of azithromycin may induce heart failure and cardiovascular mortality, potentially due to synergistic effects on QT length. We call for caution if such combination is to be used in the management of Covid-19.

**Trial registration number:** Registered with EU PAS; Reference number EUPAS34497 (http://www.encepp.eu/encepp/viewResource.htm?id=34498). The full study protocol and analysis source code can be found at https://github.com/ohdsi-studies/Covid19EstimationHydroxychloroquine.

**Funding sources:** This research received partial support from the National Institute for Health Research (NIHR) Oxford Biomedical Research Centre (BRC) and Senior Research Fellowship (DPA), US National Institutes of Health, Janssen Research & Development, IQVIA, and by a grant from the Korea Health Technology R&D Project through the Korea Health Industry Development Institute (KHIDI), funded by the Ministry of Health & Welfare, Republic of Korea [grant number: HI16C0992]. Personal funding included Versus Arthritis [21605] (JL), MRC-DTP [MR/K501256/1] (JL), MRC and FAME (APU). The European Health Data & Evidence Network has received funding from the Innovative Medicines Initiative 2 Joint Undertaking (JU) under grant agreement No 806968. The JU receives support from the European Union’s Horizon 2020 research and innovation programme and EFPIA. No funders had a direct role in this study. The views and opinions expressed are those of the authors and do not necessarily reflect those of the Clinician Scientist Award programme, NIHR, NHS or the Department of Health, England.

## INTRODUCTION

As the severe acute respiratory syndrome coronavirus 2 (SARS-CoV-2) pandemic exerts an unprecedented pressure on health care systems worldwide, there remains a paucity of evidence surrounding the safety and effectiveness of potential treatments.^1^ Several existing drugs have been postulated to be effective against SARS-CoV-2. These include conventional synthetic disease modifying anti-rheumatic drugs (csDMARDs), which are most commonly used as the first line treatment of autoimmune diseases such as rheumatoid arthritis (RA) and systematic lupus erythematosus (SLE).^2,3^ Hydroxychloroquine (HCQ) has been proposed as potential treatment options for COVID-19 based on its mechanism of action. Accumulating in the acid vesicles (endosome, Golgi vesicles, lysosomes), HCQ causes alkalinisation, leading to enzyme dysfunction and preventing endosome mediated viral entry to the cell. ^3-6^ It is also suggested in vitro that HCQ can prevent glycosylation of virus cell proteins including the ACE2 receptor, inhibiting virus entry and replication, and that similar compounds like chloroquine can specifically inhibit SARS-Cov-2.^5,7-9^ In clinical studies, the addition of HCQ has shown increased early virological response to treatment for chronic hepatitis C, and reduced viral load in patients with HIV infection, compared to placebo. ^10,11^ Treatment with HCQ also lowered IL-6 level in HIV patients, suggesting the agent may have immunosuppressive properties helpful in the prevention or treatment of cytokine storm associated with severe COVID-19 disease.^12,13^

As of 28^th^ March 2020, there are over 21 registered ongoing clinical trials and 3 prophylactic studies assessing the efficacy of hydroxychloroquine HCQ for the treatment of SARS-Cov-2.^14-20^ Early results from randomised controlled trials conducted in China have shown reduced severity and course of the disease with hydroxychloroquine HCQ, compared with placebo, without detecting serious adverse effects, although others have suggested no difference in outcome from conventional treatment.^21,22^ Of those studies that have reported more detailed results and received significant media attention, HCQ has been proposed at higher doses than used in the treatment of auto-immune disorders and alongside azithromycin (AZM), a macrolide antibiotic.^23 24^ Results from this open label observational study suggest that the combination of HCQ and azithromycin AZM might lead to a faster recovery and reductions in viral load in the treatment of COVID-19. However, many authors have criticised the study due to lack of low power, limited follow-up, confounding by indication, and lack of adherence to the allocated treatment arm.^25^ The efficacy of HCQ in combination with AZM is therefore yet to be established, but approval for compassionate use by regulators and media attention will likely lead to an increase in use of this combined therapy for the management of COVID-19 worldwide.

In preparation for our study, we systematically searched the literature (PubMed, Embase), clinical trial registries (Clinicaltrials.gov, ICTRP and Chinese Clinical Trial Registry) and preprint servers (bioRxiv and medRxiv) from inception until 27/03/2020 (Supplementary appendix section 11). No contemporary large-scale evidence was found to identify the real-world comparative safety of HCQ compared to other first line DMARDs, especially in combination with macrolide antibiotics such as AZM that are being considered for use in treating COVID-19.

Sepriano *et al*. led a systematic review to inform EULAR 2019 recommendations for the safety of RA medications, but little high-level evidence focussed on HCQ.^26^ Another recent review of the comparative risks of non-serious and serious adverse events (SAEs) associated with DMARDs predominantly focussed upon biologic therapies.^27^ There is little good high quality evidence quantifying SAEs risk in the literature with several studies suggesting no increased infection risk with any nonbiologic DMARDs, including HCQ.^28,29^ The safety profile of HCQ is described in its summary of products characteristics, with adverse drug reactions including severe cardiac disorders as QT segment prolongation that could lead to arrhythmia, myocardial arrest or cardiovascular death.^30^ Azithromycin (AZM, and macrolides in general) are known to induce cardiotoxicity when used alone, and to also increase the risk of other drugs that prolong QTc interval.^31-34^ It is therefore of utmost importance that we understand the safety implications of the proposed combination of HCQ and azithromycin AZM before this becomes standard practice in the management of COVID-19 globally.

In light of the current global pandemic, information regarding the safety of HCQ in worldwide real-world practice is vital to inform policy.^35,36^ We aimed to assess the safety of hydroxychloroquine (HCQ) alone and in combination with AZM to help guide decisions in the face of the growing COVID-19 pandemic.

## METHODS

### Study design

Two study designs were developed and executed across a multinational, distributed database network. First, new user cohort studies were used to estimate the safety of HCQ compared to sulfasalazine (SSZ), and to assess the risks associated with the addition of AZM compared to amoxicillin (AMX) amongst users of HCQ in patients with rheumatoid arthritis (RA). SSZ and AMX were chosen as active comparators as they have similar indications as the target treatments (HCQ and AZM respectively). As a secondary analysis, self-controlled case series (SCCS) was used to estimate the safety of HCQ in the wider population, including uses for non-RA indications.

### Data sources

Electronic health records and administrative claims databases from primary care and secondary care containing participants from Germany, Japan, Netherlands, Spain, the UK, and the USA were analysed in a distributed network, and are detailed in the Supplementary Appendix, Table S1.

Observational healthcare databases mapped to the Observational Medical Outcomes Partnership (OMOP) common data model collaborated in an international effort with the Observational Health Data Science and Informatics (OHDSI) community.^37,38^ De-identified or pseudonymised data were obtained from routinely collected records from clinical practice in Germany, Spain, the UK, Japan, and the USA. Studies were performed locally and no patient level data shared using the following databases: IQVIA Disease Analyser Germany EMR (ambulatory EMR from Germany); JMDC (Japanese claims); IPCI (primary care EMR from Netherlands); SIDIAP (primary care EMR from Spain); CPRD and IMDR (primary care EMRs from UK); and CCAE, Optum, MDCR, MDCD, PanTher, IQVIA OpenClaims, Veteran Affairs (VA), and IQVIA US Ambulatory EMR (USA). SCCS were conducted on a subset of these as a secondary analysis: CCAE, CPRD, Optum, MDCD, and MDCR. Rather than pooling these data assets, all analyses were conducted in a distributed network, where analysis code was sent to participating sites and only aggregate summary statistics were returned, with no sharing of patient-level data between organizations.

### Study Period and Follow-up

The study period started from 01/09/2000 and ended at the latest available date for all data sources in 2020. Follow-up for each of the cohorts started at an index date defined by the first dispensing or prescription of the target/comparator drug as described in the cohort definitions (Supplementary Table 2.1). Two periods were considered to define time-at-risk. First, for an *intention-to-treat analysis*, follow-up started one day after the index date and continued up until the first of: outcome of interest, loss to follow-up, or 30 days after the index date to resemble the likely duration of COVID-19 treatment regimens.^23^ Secondly, for an *on-treatment analysis*, follow-up started one day after the index date and continued until the earliest of: outcome of interest, loss to follow-up, or discontinuation, with an added washout time of 14 days. Continued use of a same treatment was inferred by allowing up to 90-day gaps between dispensing or prescription records.

In the HCQ versus SSZ study, the index event was defined as the first recorded dispensing or prescription of the drug in a patient’s history. For the study of HCQ combined with AZM, follow up started when the second of the two co-administered treatments was initiated while still exposed to the first treatment (e.g. when AZM started during a period of HCQ use, or when HCQ started during a period of AZM use). HCQ use was assumed to be chronic in the management of RA, and AZM was assumed an acute prescription for infection treatment, and therefore inferred persistent exposure to AZM was assessed by allowing up to 30 days between dispensing or prescription records. Cohorts of combined HCQ and amoxicillin were generated using these same rules as an active comparator.

For SCCS, periods of inferred persistent exposure to HCQ were generated by allowing up to 90-day gaps between dispensing or prescription records. Individual SCCS analyses were executed separately for each of the proposed study outcomes, including both safety events and negative control outcomes. Patients were followed for their entire observation time (e.g. from enrolment to disenrollment in each database), and incidence rates of each of the study outcomes calculated in periods of inferred persistent exposure to HCQ and non-exposure periods.

### Participants

For the new user cohorts, participants included those with a history of RA (a condition occurrence or observation indicating RA any time before or on the same day as therapy initiation), aged 18 years or over at the index event, with at least 365 days of continuous observation time prior to index event. Inclusion and start of follow-up started at the time one of the drugs of interest (HCQ, SSZ, or addition of AZM or AMX amongst users of HCQ) was initiated after a diagnosis of RA. For the SCCS study, all prevalent users of HCQ were included, regardless of RA history or indication for HCQ therapy.

Participants were identified using pre-specified code lists reviewed by a core team of clinicians, epidemiologists, vocabulary experts, and health data scientists with extensive expertise in the use of the OMOP CDM and the OHDSI tools. The code lists in the OMOP CDM used to identify participants are listed in Supplementary Table 2.2.

### Exposures, outcomes and confounders

The proposed code lists for the identification of the study population and for the study exposures were created by clinicians with experience in the management of RA using ATLAS, and reviewed by 4 clinicians and 1 epidemiologist (Supplementary Table 2.1).^39^

A total of 16 severe adverse events (SAEs) were analysed. Hospital-based events, not available in primary care records (CPRD, IMRD and SIDIAP), included gastrointestinal bleeding, acute renal failure, acute pancreatitis, myocardial infarction, stroke, transient ischaemic attack, and cardiovascular events (composite). Additionally, angina/chest pain, heart failure, cardiac arrhythmia, bradycardia, venous thromboembolism, end stage renal disease, and hepatic failure were analysed from both primary and secondary care data. Mortality outcomes were obtained only from data sources with reliable information on death date (CPRD, IMRD, IPCI, Optum, SIDIAP, VA) and cardiovascular events preceding death records (CPRD, IMRD, Optum, VA), with the former contributing to informing all-cause mortality, and the latter also used to assess to cardiovascular death. All codes for the identification of the 16 proposed study outcomes were based on a previously published paper, and are detailed in Supplementary Table 2.2.^40^ Face validity for each of the outcome cohorts was further reviewed by exploring age- and sex-specific incidence rates compared to previous clinical knowledge and/or existing literature.

Two active comparator analyses were conducted in the cohort studies: first, incident users of HCQ were compared to new users of SSZ; second, new use of AZM amongst prevalent users of HCQ was compared to incident use of AMX during ongoing HCQ use.

Exposure commenced on the first day of dispensing or prescription recorded with at least 365 days of prior observation period to increase confidence that the exposure was incident. Exposure interval gaps of ≤90 days (HCQ and SSZ) and of ≤30 days (AZM and AMX) between drug dispensing or prescription records were allowed and inferred as persistent exposure. Drug discontinuation was considered in the HCQ study if a patient switched from one study drug to another. Patients who switched from target exposure to comparator exposure, or vice versa, contributed follow-up time to the exposure cohort that they entered first, and were censored at the time of switching in the ‘on treatment’ analysis.

A list of negative control outcomes was also assessed for which there is no known causal relationship with any of the drugs of interest. These outcomes were identified using a semi-automatic process based on data extracted from literature, product labels, and spontaneous reports, and confirmed by manual review by 2 clinicians.^41^ A full list of codes used to identify negative control outcomes can be found in Supplementary Table 3, and details on covariate/confounder identification are provided in Supplementary Table 4.

### Study size

This study was undertaken using routinely collected data and all patients meeting the eligibility criteria above during the study observation period were included. No *a priori* sample calculation was performed; instead, a minimum detectable rate ratio (MDRR) was estimated for each drug-outcome pair in each of the available databases. The MDRRs for each of the databases for each drug pair-outcome analysis, as well as sample size for each of the comparisons are reported in full in an interactive web app (https://data.ohdsi.org/Covid19EstimationHydroxychloroquine/. Only analyses with 0 counts in either treatment group were excluded based on power, with all others contributing to meta-analytic estimates where applicable.

### Statistical methods

PS stratification was used as the analytical strategy to adjust for imbalance between exposure cohorts in a comparison, using a large-scale regularized logistic regression ^36^ fitted with a LASSO penalty and with the optimal hyperparameter determined through 10-fold cross validation. Baseline patient characteristics were constructed for inclusion as potentially confounding covariates.^42^ From this large set of tens of thousands of covariates, key predictors of exposure classification were selected for the propensity score. The predictor variables included were based on all observed patient characteristics and covariates available at each data source, including conditions, procedures, visits, observations and measurements. All covariates that occur in fewer than 0.1% of patients within the target and comparator cohorts were excluded prior to propensity score model fitting for computational efficiency. Patients in the target and comparator cohorts were stratified into 5 propensity score quintiles.

Plotting the propensity score distribution and assessment of covariate balance expressed as the standardized difference of the mean was undertaken for every covariate before and after propensity score adjustment. A standardized difference > 0.1 indicated a non-negligible imbalance between exposure cohorts.^43^ The target and comparator cohort were compared using a univariate Cox proportional hazards model conditioned on the propensity score strata with treatment allocation as the sole explanatory variable. Negative control outcomes analyses and empirical calibration were used to further minimise potential unresolved confounding with calibrated HRs (CalHRs) and 95% confidence intervals estimated.^44,45^

For SCCS, safety of HCQ therapy was assessed separately as a secondary analysis, regardless of indication, comparing exposed and unexposed time periods within the same individuals. The method is self-controlled in that it makes within-person comparisons of event rates during periods of hypothesized increase risk with other periods of baseline risk, with eliminates all time-invariant confounding. Because we do not compare between persons, the SCCS is robust to between-person differences, even including unmeasured differences (like genetics). However, the method is vulnerable to time-varying confounders: the time of exposure may be incomparable to the time when not exposed. To adjust for this, we included many time-varying co-variates in the models, including age, season, and other drug exposures. The effects of age and season were assumed constant within each calendar month and were modelled using bicubic splines with 5 knots. A conditional Poisson regression was used to fit the outcome model using the Cyclops package, with a hyperparameter selected through 10-fold cross-validation.^46^

Study diagnostics (power, propensity score distribution, covariate balance, empirical null distribution) were evaluated by clinicians and epidemiologists to determine which database-target-comparator-outcome-analysis variants could produce unbiased estimates. Database-target-comparator-analysis variants with zero event outcomes in the time-at-risk window or contained analyses with baseline covariate with standardized mean difference>0.1 after stratification were excluded from analysis. Study diagnostics for all database-target-comparator-outcome-analysis will be provided as part of study, regardless of which effect estimation results are unblinded. All the proposed analyses were conducted for each database separately, with estimates combined in fixed effects meta-analysis methods where I2 is <=40%. No meta-analysis was conducted where I2 for a given drug-outcome pair is >40%. Of note, when running analysis in a distributed network, it was not possible to link across datasets, and to know the extent of overlap between data.

All analytical code is available at https://github.com/ohdsi-studies/Covid19EstimationHydroxychloroquine, with study diagnostics considered prior to the unblinding of estimation results. All study diagnostics are available for exploration at https://data.ohdsi.org/Covid19EstimationHydroxychloroquine/. All statistical analyses were conducted using tools previously validated by the OHDSI community. For the cohort analysis, the CohortMethod package was used (https://ohdsi.github.io/CohortMethod/) using a large-scale propensity score (PS) constructed through the Cyclops package (https://ohdsi.github.io/Cyclops).^46^ All SCCS were run using the freely available package (https://ohdsi.github.io/SelfControlledCaseSeries/).^47^

## RESULTS

### Participants

A total of 956,374 HCQ and 310,350 SSZ users were identified, with 323,122 and 351,956 contributing to the analyses of combination therapy of HCQ with AZM compared to HCQ with AMX respectively. Participant counts in each data source are provided in Appendix S5.

Users of HCQ were more likely female (e.g. 82.0% vs 74.3% in CCAE) and less likely to have certain comorbidities like inflammatory bowel disease (e.g. prevalence of Crohn’s disease 0.6% vs 1.8% in CCAE) or psoriasis (e.g. 3.0% vs 8.9% in CCAE). All these differences were however minimised after propensity score stratification, with all reported analyses balanced on all identified confounders including socio-demographics, comorbidities and concomitant drug/s use. Similarly, users of combination HCQ+AZM differed from those of HCQ+AMX, with a prevalence of acute respiratory disease appearing higher amongst azithromycin users (62.5% vs 50.7% in CCAE). Again, propensity score methods resolved these differences, and comparison groups became balanced for all observed confounders after stratification. Detailed baseline characteristics for HCQ vs SSZ and for HCQ+AZM vs HCQ+AMX after propensity score stratification in CCAE are detailed in Table 1 for illustrative purposes, and similar tables with a more complete list of features for each included database and comparing before and after propensity score stratification are provided as Supplementary Tables 6.1.1 to 6.1.14 for HCQ vs SSZ, and Supplementary Tables 6.2.1 to 6.2.13 for HCQ+AZM vs HCQ+AMX.

**Table 1.**
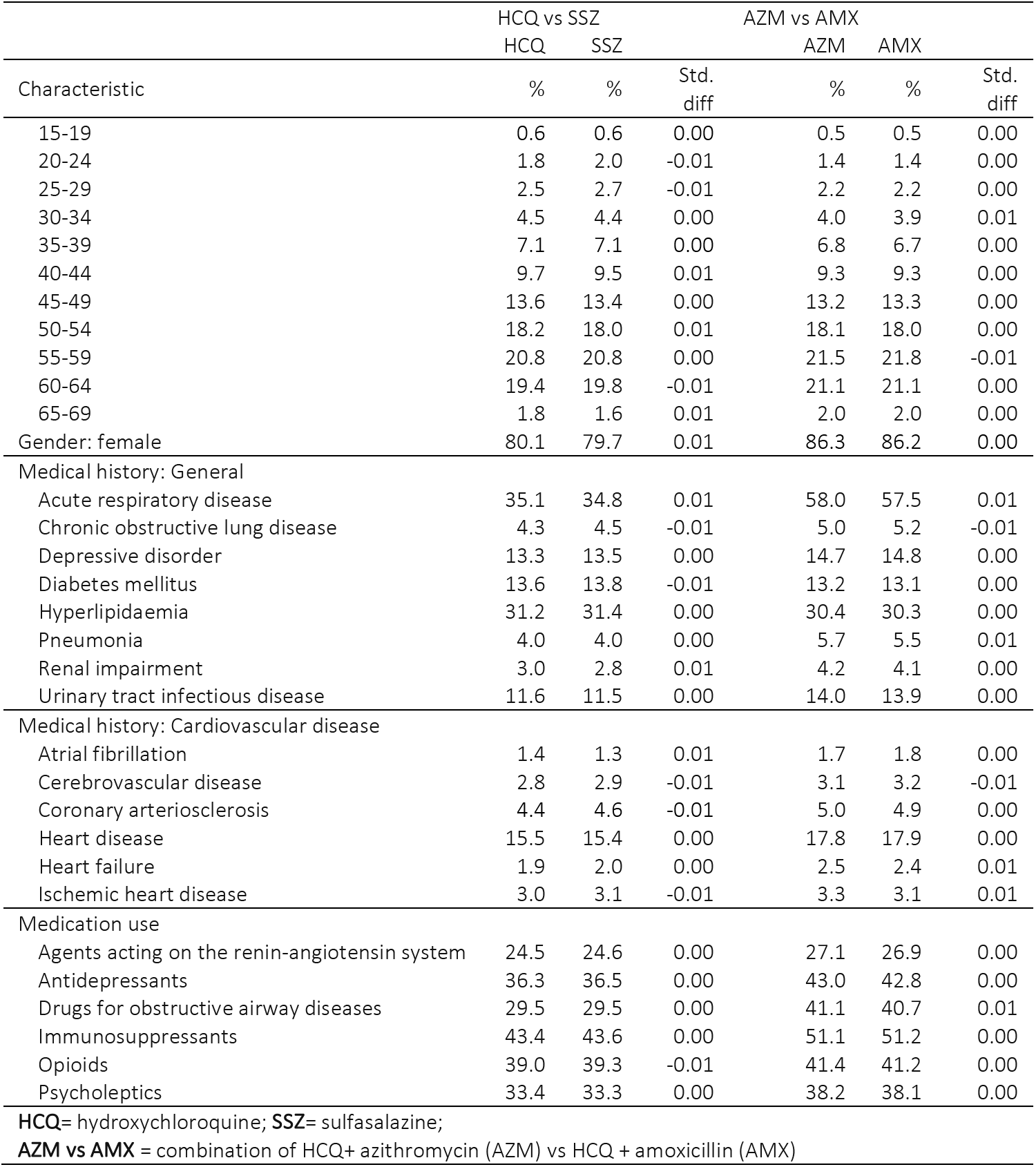
Baseline characteristics of users of HCQ compared to SSZ, and HCQ+AZM vs HCQ+AMX after propensity score stratification in CCAE

Propensity score distribution plots showing overlap between groups and figures depicting all covariate balance and empirical null distribution plots based on negative controls can be found in Supplementary Tables 9.1 to 9.14 (Evidence evaluation diagnostics), and interactive versions of these are available at https://data.ohdsi.org/Covid19EstimationHydroxychloroquine/

### Outcome Data

We report here (Table 2) on database-specific counts and rates of key outcomes (cardiovascular mortality, chest pain/angina and heart failure) observed in the proposed 30-day *intention-to-treat* analysis.

**Table 2.**
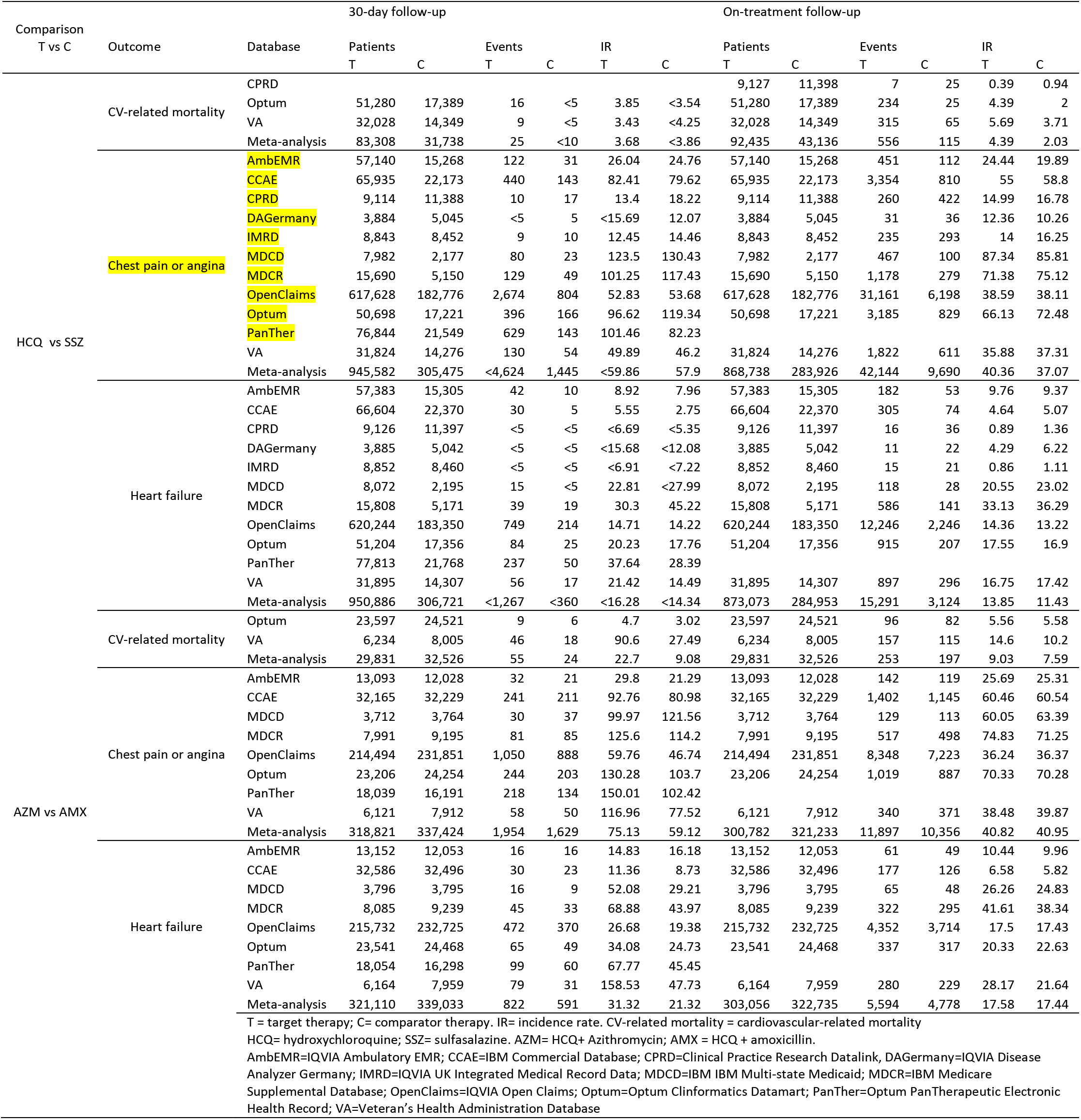
Event occurrence

Database-specific counts, incidence rates (IR) of all study outcomes stratified by drug use are detailed in full in Supplementary Table S7. Least common outcomes included bradycardia (e.g. IR 0.92/1,000 person-years (py) amongst HCQ users in CCAE) and end-stage renal disease (e.g. IR <0.92/1,000 py amongst HCQ users in CCAE), whilst most common ones were chest pain/angina (e.g. IR 82.41/1,000 py amongst HCQ users in CCAE) and composite cardiovascular events (e.g. IR 17.96/1,000 py amongst HCQ users in CCAE). As expected, most IRs appeared higher in data sources which included older populations (e.g. IR of composite cardiovascular events in HCQ users in MDCR of 91.39/1,000 py). Mortality rates ranged from 4.81/1,000 person-years in HCQ users in Optum to 17.13/1,000 py amongst HCQ users in VA, with cardiovascular-specific mortality ranging from IR 3.43/1,000 py in HCQ users in VA to <4.25/1,000 person-years in SSZ users in the same data source.

Database and outcome-specific HRs (uncalibrated as well as calibrated) are reported in full in the form of forest plots (Supplementary Figure Sections 8.1 and 8.2). None of the SAEs appeared consistently increased with the short-term use of HCQ (vs SSZ) in the *intention-to-treat* analyses (Figure 1), with meta-analytic calibrated HRs (CalHRs and 95%CI) ranging from 0.67 (0.45-1.01) for hepatic failure to 1.35 (0.51-3.63) for cardiovascular mortality (Figure 2).

**Figure 1.**
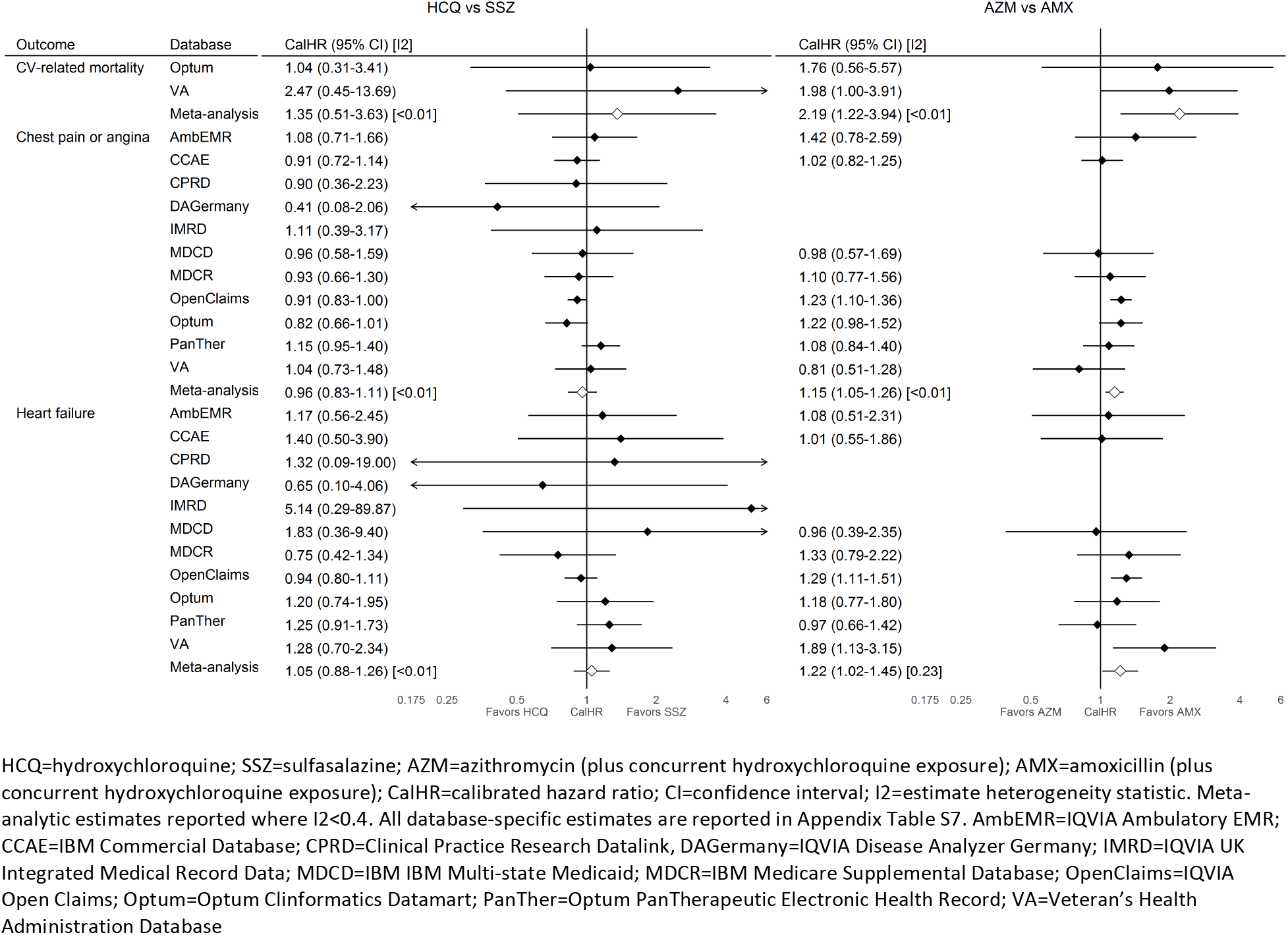
Source-specific and meta-analytic cardiovascular risk estimates for hydroxychloroquine vs sulfasalazine and azithromycin vs amoxicillin new users during 30-day follow-up

**Figure 2.**
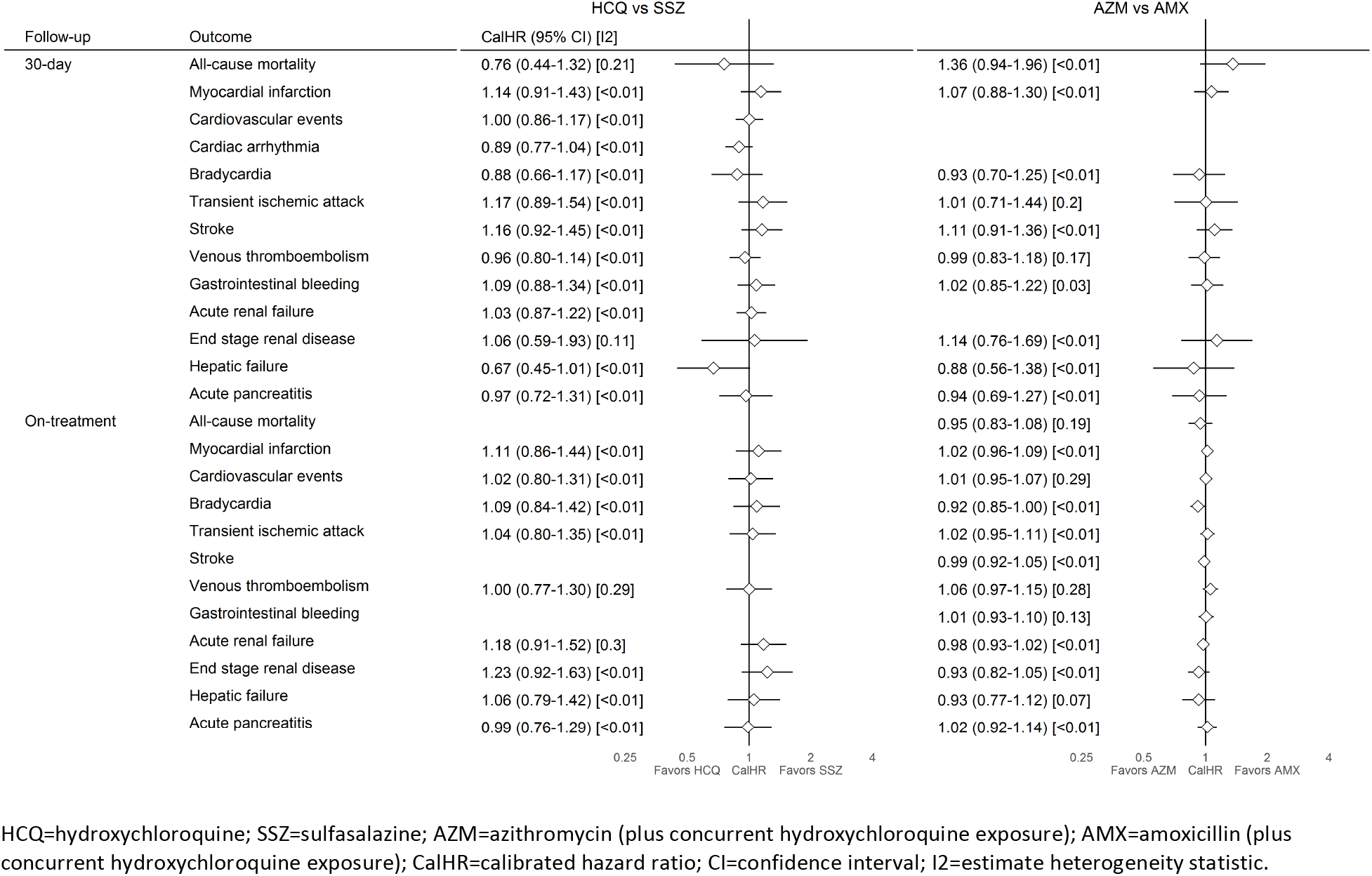
Meta-analytic risk estimates for hydroxychloroquine vs sulfasalazine and azithromycin vs amoxicillin new users during on-treatment during 30-day and on-treatment follow-up

Consistent findings were seen with the long-term (*on treatment*) use of HCQ vs SSZ (Figure 3), with the exception of cardiovascular mortality, which appeared inconsistent in the available databases, but overall increased in the HCQ group when meta-analysed: pooled CalHR 1.65 (1.12-2.44).

**Figure 3.**
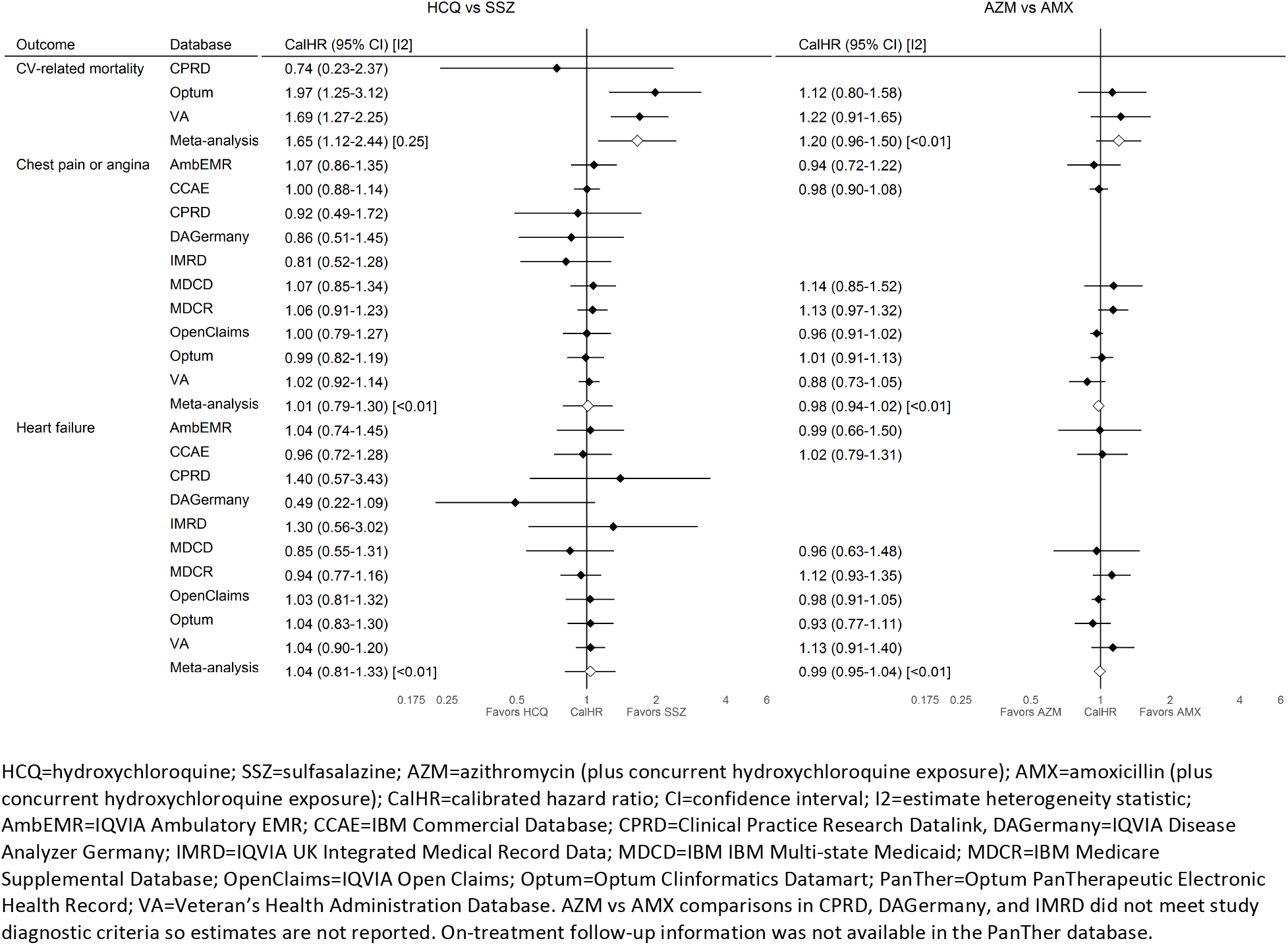
Source-specific and meta-analytic cardiovascular risk estimates for hydroxychloroquine vs sulfasalazine and azithromycin vs amoxicillin new users during on-treatment follow-up

Similar results were obtained in SCCS analyses, which looked at the effect of HCQ use (on-vs off-treatment) on all outcomes except mortality regardless of indication, and therefore included non-RA patients (Tables S10.1 to 10.6 for database-specific results).

All the obtained database- and outcome-specific calHRs for the association between short-term (1 month) use HCQ+AZM vs HCQ+AMX are depicted in the form of Forest plots in Supplementary Figure Sections 8.1 and 8.2. Three SAEs appeared increased with the short-term (30-day fixed follow-up) use of HCQ+AZM: chest pain/angina (meta-analytic CalHR 1.15 (1.05-1.26), heart failure (meta-analytic CalHR 1.22 (1.02-1.45)), and cardiovascular mortality (meta-analytic CalHR 2.19 (1.22-3.94) (Figure 1).

## DISCUSSION

Despite a lack of evidence on efficacy, HCQ and HCQ+AZM have become the most popular treatment/s for COVID-19. This is the largest ever analysis of the safety of such treatments worldwide, examining over 900,000 HCQ and more than 300,000 HCQ+AZM users respectively.

The results on the risk of SAEs associated with short-term (1 month) HCQ treatment as proposed for COVID-19 therapy are reassuring, with no excess risk of any of the considered safety outcomes compared to an equivalent therapy (SSZ). However, long-term treatment with HCQ as used for RA is associated with a 65% increase in cardiovascular mortality.

Worryingly, significant risks are identified for combination users of HCQ+AZM even in the short-term as proposed for COVID19 management, with a 15-20% increased risk of angina/chest pain and heart failure, and a two-fold risk of cardiovascular mortality in the first month of treatment.

A systematic review of the cardiac side effects of chloroquine and HCQ identified 86 articles reporting short series or individual cases.^39^ In the 127 included patients, cardiac side effects occurred in mainly women (65.4%) who had a median age of 56 years. Conduction disorders were the main side effect reported (85%), with heart failure (26.8%), ventricular hypertrophy (22%), hypokinesia (9.4%), valvular dysfunction (7.1%) and pulmonary arterial hypertension (3.9%) being the other reported side effects. When drugs were withdrawn, 44.9% of patients recovered normal cardiac function; 12.9% sustained irreversible damage, and 30% died. It should be noted that cardiac toxicity was induced by a high cumulative dose of chloroquine or HCQ in most patients, although some studies identified by this systematic review mentioned complications even in patients with a low cumulative dose. Furthermore, interrogation of the Food and Drug Administration’s adverse event reporting database FAERS from 2004-2019 Q4 saw 357 adverse events reported.^48^ 20% of the events reported were cardiac, with the median age of patients included being 39, and a male to female ratio of 0.60. The cardiovascular SAEs reported appear similar to those included in the review by Chatre *et al*., with complete AV block 1.8%; cardiac arrest 1.8% ventricular fibrillation 1.09%, cardiogenic shock 0.6%; heart failure 1.4%; cardiomyopathy 1.6% reported as the most likely cardiovascular SAEs.

Our results suggest that long-term use of HCQ leads to an increased risk of cardiovascular mortality, with no observable excess risk of major cardiovascular events or diagnosed bradycardia. Considering the current evidence, this may relate to cumulative effects of HCQ leading to an increased risk of QT lengthening or relate to the moderately increased risk of angina and heart failure seen. However, as the strong association observed with cardiovascular death is not observed with diagnosed arrhythmia or bradycardia in this study, sudden cardiovascular death here is more likely due to QT lengthening and undetected and/or sudden torsade-de-pointes. Although long-term treatment with HCQ is not expected for the management of COVID-19, some research suggests that higher doses as prescribed for COVID-19 can, even in the short-term, lead to equivalent side effects given the long half-life of HCQ.^49^

QT lengthening is a known effect of all macrolides including AZM and physicians already use caution when prescribing macrolides concurrently with other medications that can also increase the QT interval.^32-34^ In this study, the elevated risk of cardiovascular death with combined HCQ +AZM therapy may arise through their synergistic effects of inducing lethal arrhythmia.

As with all observational data, this study is limited by its ability to appropriately identify exposure and outcome. Due to the nature of sudden cardiac death, capturing the true cause of cardiovascular related mortality is difficult. We therefore have explored cardiovascular related outcomes other than mortality to determine if deterioration in these pathophysiological processes led to increased mortality. Since this is not seen, and sudden cardiac death in association with prolonged QT interval is described in the literature, our conclusions are drawn from these assumptions. It should be acknowledged that misclassification can occur due to non-adherence or non-compliance with exposure medication, and incomplete lack of recording of SAEs may lead to underestimation of these outcomes.

Another potential limitation in this study is the potential for patients to be included in more than one dataset in the US. Whilst we ran meta-analysis, which assume populations are independent, we wish to highlight we are likely to under-estimate variance in our meta-analytic estimates.

The comparative new user cohort studies are anchored in patients using HCQ for RA, who therefore are likely to be using HCQ at a lower dose than is currently being proposed for use in the treatment of COVID-19. We have taken into consideration that patients with RA taking HCQ may also have further auto-immune conditions such as systemic lupus erythematosus (SLE) and therefore generate the potential for confounding by indication.^50^ We therefore ensured that when investigating covariate balance after propensity score stratification and matching and before unblinding study results, that we did not see unbalanced proportions of patients with a diagnosis of SLE between the groups. Negative control outcome analyses also did not identify any residual unobserved confounding in the PS analysis. Whilst patients with RA may have greater levels of comorbidities than the general population, the age and demographic profile of patients developing cardiovascular complications described in both the systematic review and FAERS database suggests that complications are not only restricted to those with multimorbidity.^48^ However, absolute risk in our study should be interpreted cautiously since patients with RA are likely different from those with COVID-19.

As the world awaits the results of clinical trials for the anti-viral efficacy of HCQ in the treatment of SARS-Cov2 infection, this large scale, international real-world data network study enables us to consider the safety of the most popular drugs under consideration. HCQ appears to be largely safe in both direct and comparative analysis for short term use, but when used in combination with AZM this therapy carries double the risk of cardiovascular death in patients with RA. Whereas we used the collective experience of a million patients to build our confidence in the evidence around the safety profile, the current evidence around efficacy of HCQ+AZI in the treatment of covid-19 is quite limited and controversial.

## Data Availability

This study was conducted as a distributed database network analysis. To protect patient privacy, all patient-level data were maintained securely behind institutional firewalls. Analysis code was downloaded and executed by each participating data partner, which generated only aggregate summary statistics (cohort counts, model coefficients) which were then centralized and synthesized in preparation of this manuscript. All aggregate summary statistics produced have been made publicly available at: https://data.ohdsi.org/Covid19EstimationHydroxychloroquine/.

https://github.com/ohdsi-studies/Covid19EstimationHydroxychloroquine

https://github.com/ohdsi-studies/Covid19DrugRepurposing

## ETHICAL APPROVAL

All data partners received IRB approval or waiver in accordance to their institutional governance guidelines.

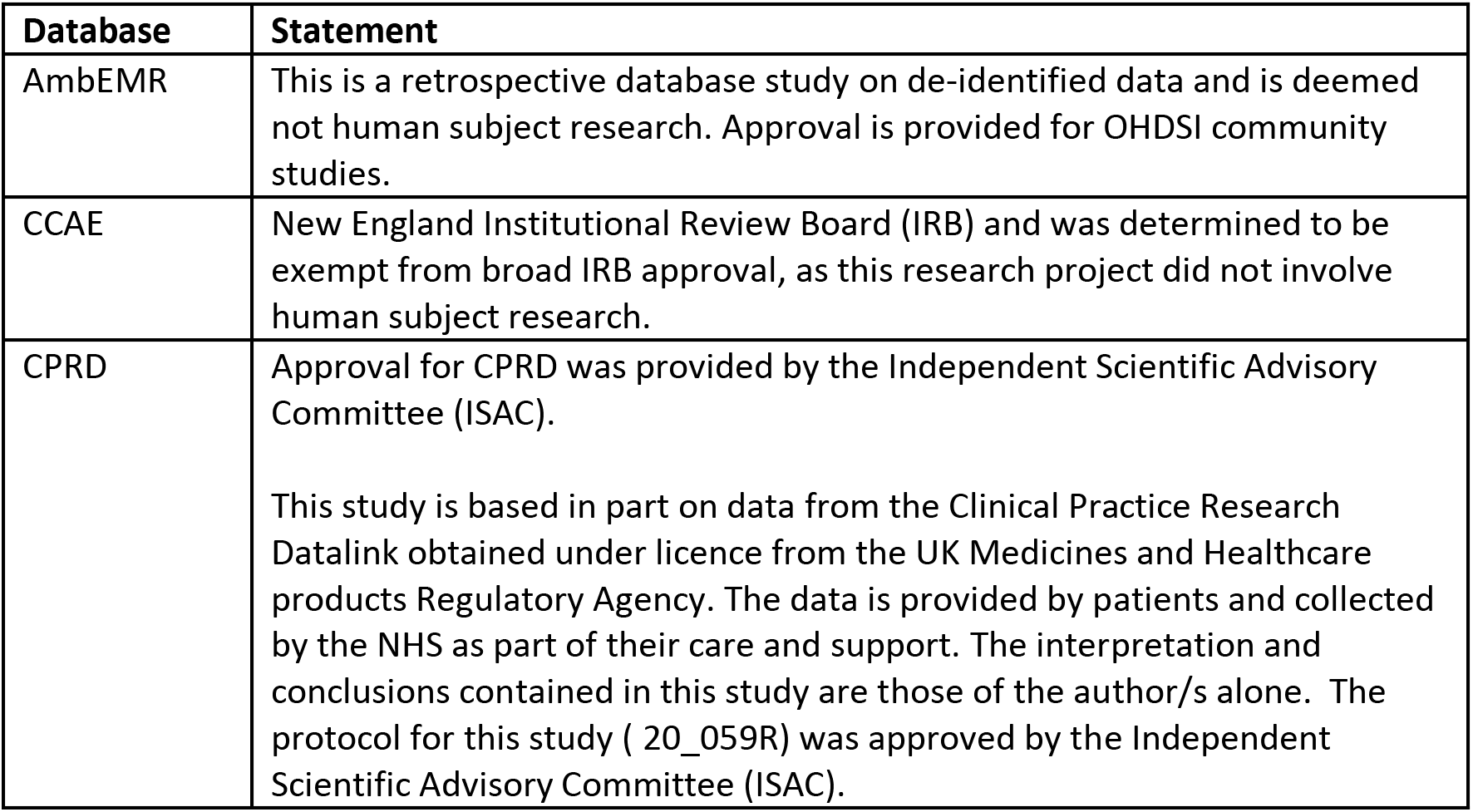
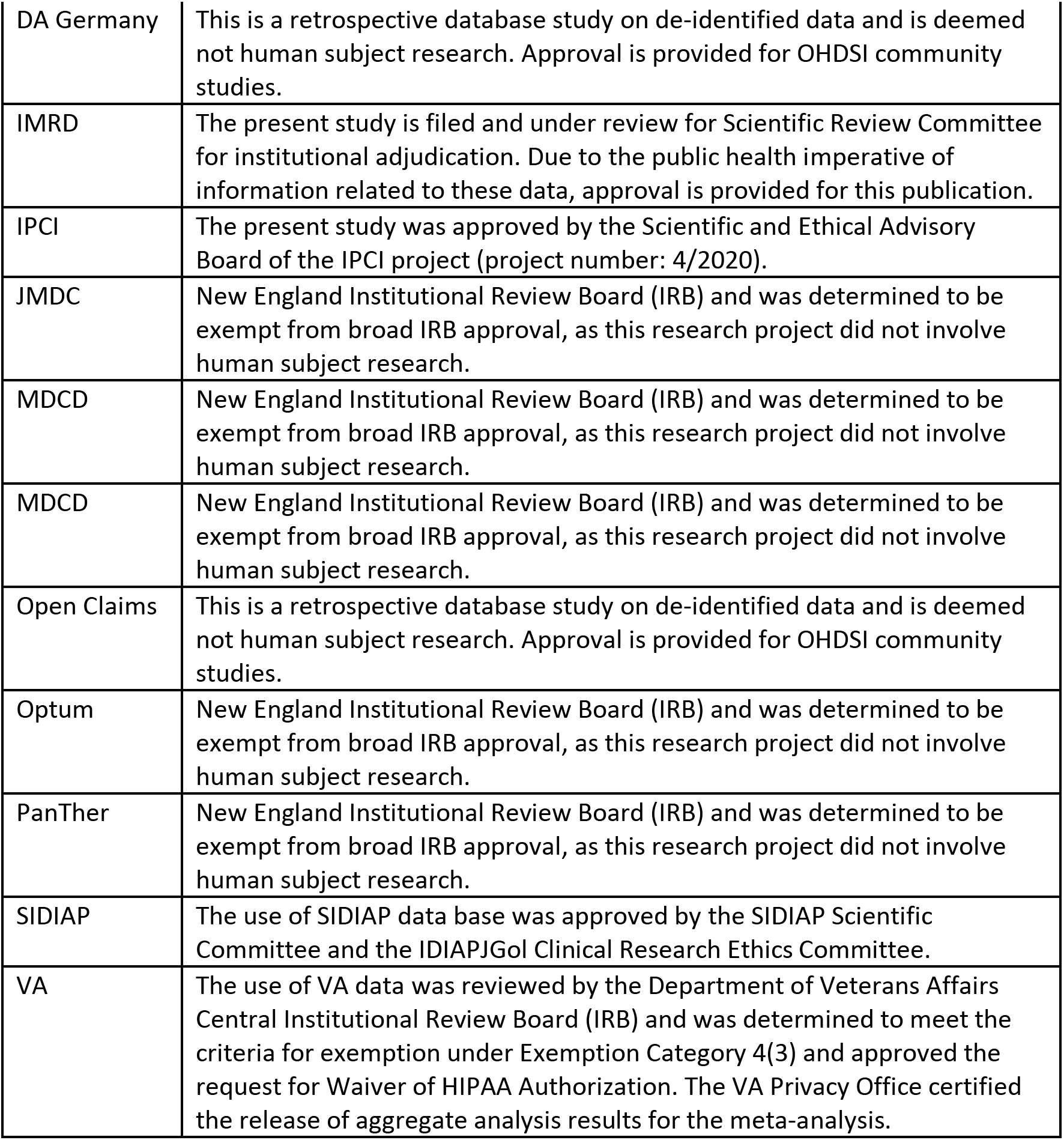

## DECLARATION OF INTERESTS

All authors have completed the ICMJE uniform disclosure form at www.icmje.org/coidisclosure.pdf

## ACKNOWLEDGEMENTS

Catherine Hartley and Eli Harriss, Bodleian Health Care Libraries, University of Oxford, Nigel Hughes; Runsheng Wang, Zeshan Ghosh, Liliana Ciobanu and Michael Kallfelz. Finally, we acknowledge the tremendous work and dedication of the 350 participants from 30 nations in the March 2020 OHDSI COVID-19 Virtual Study-a-thon (https://www.ohdsi.org/covid-19-updates/), without whom this study could not have been realized.

## Notes

### Competing Interest Statement

all authors have completed an ICJME form

### Clinical Trial

Registered with EU PAS; Reference number EUPAS34497 (http://www.encepp.eu/encepp/viewResource.htm?id=34498). The full study protocol and analysis source code can be found at https://github.com/ohdsi-studies/Covid19EstimationHydroxychloroquine.

### Clinical Protocols

http://www.encepp.eu/encepp/viewResource.htm?id=34498

https://github.com/ohdsi-studies/Covid19EstimationHydroxychloroquine.

## REFERENCES

1. WHO. Report of the WHO-China Joint Mission on Coronavirus Disease 2019 (COVID-19). Geneva: WHO; 2020.

2. Smolen JS, Landewe R, Breedveld FC, et al. EULAR recommendations for the management of rheumatoid arthritis with synthetic and biological disease-modifying antirheumatic drugs: 2013 update. Ann Rheum Dis 2014;73:492–509.

3. Colson P, Rolain J-M, Lagier J-C, Brouqui P, Raoult D. Chloroquine and hydroxychloroquine as available weapons to fight COVID-19. International journal of antimicrobial agents 2020:105932-.

4. Savarino A, Boelaert JR, Cassone A, Majori G, Cauda R. Effects of chloroquine on viral infections: An old drug against today’s diseases? Lancet Infectious Diseases 2003;3:722–7.

5. Fedson DS. Confronting an influenza pandemic with inexpensive generic agents: can it be done? The Lancet Infectious Diseases 2008;8:571–6.

6. Vigerust DJ, Shepherd VL. Virus glycosylation: role in virulence and immune interactions. Trends in Microbiology 2007;15:211–8.

7. Devaux CA, Rolain J-M, Colson P, Raoult D. New insights on the antiviral effects of chloroquine against coronavirus: what to expect for COVID-19? International journal of antimicrobial agents 2020:105938-.

8. Savarino A, Di Trani L, Donatelli I, Cauda R, Cassone A. New insights into the antiviral effects of chloroquine. The Lancet Infectious diseases 2006;6:67–9.

9. Wang M, Cao R, Zhang L, et al. Remdesivir and chloroquine effectively inhibit the recently emerged novel coronavirus (2019-nCoV) in vitro. Cell Res 2020;30:269–71.

10. Helal GK, Gad MA, Abd-Ellah MF, Eid MS. Hydroxychloroquine augments early virological response to pegylated interferon plus ribavirin in genotype-4 chronic hepatitis C patients. Journal of medical virology 2016;88:2170–8.

11. Sperber K, Louie M, Kraus T, et al. Hydroxychloroquine treatment of patients with human immunodeficiency virus type 1. Clin Ther 1995;17:622–36.

12. Sperber K, Chiang G, Chen H, et al. Comparison of hydroxychloroquine with zidovudine in asymptomatic patients infected with human immunodeficiency virus type 1. Clin Ther 1997;19:913–23.

13. Mehta P, McAuley DF, Brown M, Sanchez E, Tattersall RS, Manson JJ. COVID-19: consider cytokine storm syndromes and immunosuppression. Lancet 2020;395:1033–4.

14. ChiCTR2000029542. Study for the efficacy of chloroquine in patients with novel coronavirus pneumonia (COVID-19). 2020.

15. ChiCTR2000029740. Efficacy of therapeutic effects of hydroxycholoroquine in novel coronavirus pneumonia (COVID-19) patients(randomized open-label control clinical trial). 2020.

16. ChiCTR2000029740, The First Hospital of Peking University Y. Efficacy of therapeutic effects of hydroxycholoroquine in novel coronavirus pneumonia (COVID-19) patients(randomized open-label control clinical trial). 2020.

17. ChiCTR2000029559, Renmin Hospital of Wuhan University N. Therapeutic effect of hydroxychloroquine on novel coronavirus pneumonia (COVID-19). 2020.

18. ChiCTR2000029559. Therapeutic effect of hydroxychloroquine on novel coronavirus pneumonia (COVID-19). 2020.

19. ChiCTR2000029741, The Fifth Affiliated Hospital Sun Yat-Sen University Y. Efficacy of Chloroquine and Lopinavir/ Ritonavir in mild/general novel coronavirus (CoVID-19) infections: a prospective, open-label, multicenter randomized controlled clinical study. 2020.

20. ChiCTR2000029609, The Fifth Affiliated Hospital of Sun Yat-Sen University Y. A prospective, open-label, multiple-center study for the efficacy of chloroquine phosphate in patients with novel coronavirus pneumonia (COVID-19). 2020.

21. Gao J, Tian Z, Yang X. Breakthrough: Chloroquine phosphate has shown apparent efficacy in treatment of COVID-19 associated pneumonia in clinical studies. Biosci Trends 2020;14:72–3.

22. Efficacy and Safety of Hydroxychloroquine for Treatment of Pneumonia Caused by 2019-nCoV (HC-nCoV). 2020. at https://clinicaltrials.gov/show/NCT04261517;http://subject.med.wanfangdata.com.cn/UpLoad/Files/202003/43f8625d4dc74e42bbcf24795de1c77c.pdf.)

23. Gautret P, Lagier JC, Parola P, et al. Hydroxychloroquine and Azithromycin as a treatment of COVID-19: preliminary results of an open-label non-randomized clinical trial. medRxiv 2020:2020.03.16.20037135.

24. Paton NI, Lee L, Xu Y, et al. Chloroquine for influenza prevention: a randomised, double-blind, placebo controlled trial. Lancet Infect Dis 2011;11:677–83.

25. Molina JM, Delaugerre C, Goff JL, et al. No Evidence of Rapid Antiviral Clearance or Clinical Benefit with the Combination of Hydroxychloroquine and Azithromycin in Patients with Severe COVID-19 Infection. Med Mal Infect 2020.

26. Sepriano A, Kerschbaumer A, Smolen JS, et al. Safety of synthetic and biological DMARDs: a systematic literature review informing the 2019 update of the EULAR recommendations for the management of rheumatoid arthritis. Ann Rheum Dis 2020.

27. Costello R, David T, Jani M. Impact of Adverse Events Associated With Medications in the Treatment and Prevention of Rheumatoid Arthritis. Clin Ther 2019;41:1376–96.

28. Lacaille D, Guh DP, Abrahamowicz M, Anis AH, Esdaile JM. Use of nonbiologic disease-modifying antirheumatic drugs and risk of infection in patients with rheumatoid arthritis. Arthritis Rheum 2008;59:1074–81.

29. Salliot C, van der Heijde D. Long-term safety of methotrexate monotherapy in patients with rheumatoid arthritis: a systematic literature research. Ann Rheum Dis 2009;68:1100–4.

30. FDA. Plaquenil Hydroxychloroquine Sulfate drug safety In: FDA, ed. Bethseda2006.

31. Guo D, Cai Y, Chai D, Liang B, Bai N, Wang R. The cardiotoxicity of macrolides: a systematic review. Pharmazie 2010;65:631–40.

32. Ray WA, Murray KT, Hall K, Arbogast PG, Stein CM. Azithromycin and the risk of cardiovascular death. N Engl J Med 2012;366:1881–90.

33. Fossa AA, Wisialowski T, Duncan JN, Deng S, Dunne M. Azithromycin/chloroquine combination does not increase cardiac instability despite an increase in monophasic action potential duration in the anesthetized guinea pig. Am J Trop Med Hyg 2007;77:929–38.

34. Lu ZK, Yuan J, Li M, et al. Cardiac risks associated with antibiotics: azithromycin and levofloxacin. Expert Opin Drug Saf 2015;14:295–303.

35. EMA. COVID-19: chloroquine and hydroxychloroquine only to be used in clinical trials or emergency use programmes. Amsterdam2020.

36. FDA. Request for Emergency Use Authorization For Use of Chloroquine Phosphate or Hydroxychloroquine Sulfate Supplied From the Strategic National Stockpile for Treatment of 2019 Coronavirus Disease. In: Administration UFaD, ed. Bethseda, USA 2020.

37. Observational Health Data Sciences and Informatics. 2020. at https://www.ohdsi.org [last accessed 24.03.2020].)

38. Data standardisation. at https://www.ohdsi.org/data-standardization/ [Last accessed 24.03.2020].)

39. OHDSI. The Book of OHDSI 2020.

40. Suchard MA, Schuemie MJ, Krumholz HM, et al. Comprehensive comparative effectiveness and safety of first-line antihypertensive drug classes: a systematic, multinational, large-scale analysis. Lancet 2019;394:1816–26.

41. Voss EA, Boyce RD, Ryan PB, van der Lei J, Rijnbeek PR, Schuemie MJ. Accuracy of an automated knowledge base for identifying drug adverse reactions. J Biomed Inform 2017;66:72–81.

42. Tian Y, Schuemie MJ, Suchard MA. Evaluating large-scale propensity score performance through real-world and synthetic data experiments. Int J Epidemiol 2018;47:2005–14.

43. Austin PC. Balance diagnostics for comparing the distribution of baseline covariates between treatment groups in propensity-score matched samples. Stat Med 2009;28:3083–107.

44. Schuemie MJ, Hripcsak G, Ryan PB, Madigan D, Suchard MA. Robust empirical calibration of p-values using observational data. Stat Med 2016;35:3883–8.

45. Schuemie MJ, Ryan PB, DuMouchel W, Suchard MA, Madigan D. Interpreting observational studies: why empirical calibration is needed to correct p-values. Stat Med 2014;33:209–18.

46. Suchard MA, Simpson SE, Zorych I, Ryan P, Madigan D. Massive parallelization of serial inference algorithms for a complex generalized linear model. ACM Trans Model Comput Simul 2013;23.

47. Simpson SE, Madigan D, Zorych I, Schuemie MJ, Ryan PB, Suchard MA. Multiple self-controlled case series for large-scale longitudinal observational databases. Biometrics 2013;69:893–902.

48. Luo MH, Q. Guirong, X. Wu, F. Wu B. Xu, T. Data Mining and Safety Analysis of Drugs for Novel Coronavirus Pneumonia Treatment based on FAERS: Chloroquine Phosphate Herald of Medicine (Yi Yao Dao Bao) 2020;2020–02-29 online first:1-14.

49. Chatre C, Roubille F, Vernhet H, Jorgensen C, Pers YM. Cardiac Complications Attributed to Chloroquine and Hydroxychloroquine: A Systematic Review of the Literature. Drug Saf 2018;41:919–31.

50. McGhie TK, Harvey P, Su J, Anderson N, Tomlinson G, Touma Z. Electrocardiogram abnormalities related to anti-malarials in systemic lupus erythematosus. Clin Exp Rheumatol 2018;36:545–51.

